# MAP3K7 novel variants in syndromic 46,XY DSD

**DOI:** 10.64898/2026.05.05.26352427

**Authors:** Thanh Nha Uyen Le, Shirin Moradi Fard, Alejandra P Reyes, Ngoc Can Thi Bich, Adriana Tavares Gomes, Marilyn C. Jones, Dung Vu Chi, Vincent R Harley

## Abstract

Mutations in *MAP3K7* are responsible for two distinct syndromes Cardiospondylocarpofacial (CSCF) and Frontometaphyseal dysplasia 2 (FMD2). Both are characterized by skeletal malformations, facial dysmorphisms, hearing loss, and mild intellectual disability. While cardiac defects are predominant in CSCF, keloid scar is a distinct feature in FMD2. Problem with gonadal development and disorders of sexual development (DSD) have not been previously chracterized.

Here we report three syndromic cases of 46,XY DSD with CSCF or FMD2, each carrying a novel heterozygous missense variants in *MAP3K7* (NM_145331.3:c.250G>A; p.V84M, NM_145331.3:c.195A>G; p.I65M, and NM_145331.3: c.574A>G; p.S192G). The DSD phenotypes include cryptorchidism, micropenis, small testis, and hypospadias. In silico tools predict all three variants are deleterious. All three *MAP3K7* variants occur in the kinase domain at highly conservative positions among mammals. *MAP3K7* is highly expressed in human fetal Sertoli cells. *MAP3K7* knock-out in HEK293T cells led to downregulation of *GATA4* and *FOG2* expression by RNA-Seq. Like *MAP3K1, MAP3K7* phosphorylated p38 while all three *MAP3K7* variants did not alter phosphorylated p38 compared to wildtype in HEK293T^MAP3K7-/-^ cells. Two *MAP3K7* missense mutants (p.V84M and p.I65M) ectopically activate ovarian beta catenin/ Wnt signalling in TOPFLASH assays. Our data suggest that *MAP3K7* contributes to male sex differentiation by increasing expression of pro-testis genes *GATA4* and *FOG2* in HEK293T^MAP3K7-/-^ cells and antagonizing pro-ovarian beta-catenin signalling, and that one or more of these activities were likely affected in 3 cases of 46,XY DSD with CSCF/FMD2 during sex development.

## Introduction

Disorders/ Differences of Sex Development (DSD) encompass a spectrum of congenital conditions where anatomical, sex chromosomal or sex hormonal are atypical [1]. DSD phenotypes range from mild manifestations such as cryptorchidism, hypospadias, or clitoromegaly to complete gonadal dysgenesis, which is the most complicated and least understood DSD subtype [2, 3]. DSD is classified into three categories: sex chromosome DSD, 46,XY DSD and 46,XX DSD based on the Chicago consensus 2006 [1]. While the diagnosis of sex chromosome DSD is straightforward, the molecular diagnosis for 46,XX and 46,XY DSD remain low, with almost half of cases having uninformative genetic tests, suggesting that novel DSD genes remain to be identified [2, 4, 5].

Mitogen-activated protein kinase kinase kinase 7 (*MAP3K7*) gene encodes for Transforming growth factor (TGF)-β-activated Kinase 1 (TAK1), a crucial serine/threonine kinase, and central mediator of immune response, cell survival, inflammation, and apoptosis [6–8]. Germline variants in *MAP3K7* have been associated with two autosomal dominant conditions: Cardiospondylocarpofacial (CSCF; MIM# 157800) via loss of function and Frontometaphyseal dysplasia type 2 (FMD2; MIM# 617137) via gain of function [9–11]. CSCF is characterized by growth retardation, dysmorphic facial features (e.g. prominent forehead, hypertelorism, downslanting palpebral fissures etc.), brachydactyly with carpal-tarsal fusion and extensive posterior cervical vertebral synostosis, cardiac anomalies, and deafness [9, 10]. FMD2 is a progressive skeletal dysplasia characterized by sclerosis of the skull, joint contracture, scoliosis, camptodactyly, facial dysmorphisms and keloid scars [11–13]. Many clinical manifestations of CSCF and FMD2 overlap such as skeletal deformities, facial dysmorphisms and hearing loss. While cardiac defects occur predominantly in CSCF, keloid scar is a distinct feature in FMD2. Occasionally in both syndromes mild ID occurs [9, 11, 12].

Approximately 50 patients have been reported with CSCF or FMD2 syndromes carrying *MAP3K7* variants [9, 10, 12–14]. Approximately 30 *MAP3K7* variants have been identified as pathogenic, and most are *de novo*. There is genotype – phenotype correlation among affected individuals with *MAP3K7* variants [9, 12]. However, the association between *MAP3K7* and DSD had not been investigated. Here, we describe three new cases diagnosed with CSCF or FMD2 presenting with 46,XY DSD and provide the functional evidence suggesting *MAP3K7* is involved in human male sex development.

## Materials and Methods

### Patients

Three patients presented with syndromic 46,XY DSD including probands referred by clinicians, with the third case identified via GeneMatcher (https://genematcher.org/). Clinical information, family history, hormonal profile and imaging test results were collected. Written informed consents were obtained for publication of genetic and clinical data, as well as clinical photographs. The study was conducted in aligned with approved human ethics by Monash Health Ethics Committee.

### Whole exome sequencing (WES)

Extracted genomic DNA samples from peripheral blood of affected individuals were sent to Illumina (Mexican patient) or accredited clinical diagnostic labs (the other two patients) for WES. Library preparation was performed using Illumina DNA Prep with Exome 2.5 Enrichment kit and paired-end sequencing was performed on the DNBSEQ™ platform. Raw reads were processed to remove adapters and low-quality bases, generating clean data encoded in Phred+33 format. Sequencing data were assessed with FastQC, and high-quality reads were aligned to the Ensembl Genome browser human genome assembly (GRCh38) with the Burrows-Wheeler Aligner (BWA-MEM) [15, 16]. Post-alignment quality metrics were evaluated with Qualimap. GATK was used for variant calling [17], followed by normalization with bcftools and annotation with VEP [18]. For downstream analysis, a minor allele frequency (MAF) threshold of <0.001 was applied using population databases such as gnomAD v4.1.0, and variants were further integrated into a unified database using GEMINI, which incorporates diverse annotation sources (e.g., dbSNP, ENCODE, UCSC, ClinVar, KEGG) to support interpretation and classification based on inheritance patterns. Variants were additionally evaluated using in-silico prediction tools including CADD, REVEL, and SpliceAI (Δscore > 0.2) [19–21], and prioritized according to allele frequency, predicted pathogenicity, inheritance pattern, and relevance to DSD genes. Variant classification was performed according to the [22].

### Computational analysis to predict variant effect and protein structure

Variant effect was predicted by using in silico tools such as REVEL, AlphaMissense, CADD score. Mutant-I (https://folding.biofold.org/cgi-bin/i-mutant2.0.cgi) was used to predict the variant impact on MAP3K7 protein stability.

Evolutionary conservation of the MAP3K7 region including V50del, V84M, Y113D, I65M, and S192G amino acids was explored with protein sequence alignment generated by Clustal Omega (https://www.ebi.ac.uk/Tools/msa/clustalo/) and compared with data provided by UCSC Database (https://genome.ucsc.edu).

### Mutagenesis and generation of expression construct

p3xFLAG-CMV-MAP3K7 WT expression construct was kindly provided by Prof. Stephen Robertson at Otago University, New Zealand. Site-direct mutagenesis was performed using the Agilent QuikChange II kit (Agilent, cat# 200523, USA) with our own-designed primers (Supplemental Table 1) following the manufacturer’s protocol. Mutated plasmids were subcloned using DH5alpha competent cells. The mutated MAP3K7 sequences were confirmed by Sanger sequencing and subjected to large scale DNA isolation for downstream experimental assays using Genopure Plasmid Midi kit (Roche, cat# 03143414001, Germany).

**Table 1:** Variant effect prediction of three novel missenses identified in our cases.

| Variant | REVEL score | AlphaMissense score | CADD score | gnomAD v4 & ClinVar |
| --- | --- | --- | --- | --- |
| V84M | 0.423 | Pathogenic | 32 | Absent |
| I65M | 0.248 | Pathogenic | 23.8 | Absent |
| S192G | 0.604 | Pathogenic | 29 | Absent |

### V84M knock in and MAP3K7 knock out by CRISPR

HEK293T cells were cultured and maintained in DMEM + GlutaMax (GIBCO, cat#10566-016) supplemented with 10% FBS and 1% antibiotic and anti-fungus. Cells were harvested and split once they reached 90-100% confluency.

To generate HEK293T^MAP3K7-/-^ or KO cells, guide RNA (gRNA) targeting exon 5 nearby V84M position of *MAP3K7* was designed using Custom Alt-R™ CRISPR-Cas9 guide RNA tool from IDT (https://sg.idtdna.com/site/order/designtool/index/CRISPR_CUSTOM). A pRP[CRISPR]-EGFP/Puro-hCas9-U6>{MAP3K7 gRNA1} construct expressing designed *MAP3K7* gRNA, Cas9 protein and eGFP marker was purchased from Vector Builder, Illinois, USA. HEK293T cells were seed in 6 well plate at 400 000 cell/ well density 16 hours before transfection to reach 70-80% confluency at tranfection. 1µg of pRP[CRISPR]-EGFP/Puro-hCas9-U6>{MAP3K7 gRNA1} plasmid was transfected in HEK293t cells using Lipofectamine 2000 (Invitrogen, cat# L3000015).

To generate HEK293T^MAP3K7V84M^ or KI cells, a similar experiment was conducted. A homology-directed repair (HDR) donor sequence carrying V84M variant was designed using Alt-R™ CRISPR HDR Design Tool from IDT (https://sg.idtdna.com/pages/tools/alt-r-crispr-hdr-design-tool). HDR ssDNA sequence was synthesized by IDT. 0.5µg HDR ssDNA was co-transfected with 1µg of pRP[CRISPR]-EGFP/Puro-hCas9-U6>{MAP3K7 gRNA1} plasmid into HEK293T cells.

48 hours post transfection, transfected cells expressing eGFP were sorted using FACs Aria Fusion system. Next, sorted cells were diluted into single cell per well for 96-well plate. Single expanding colony will be subjected to Sanger sequencing and Western blot (WB) to confirm whether successfully knock out MAP3K7 or knock in V84M.

### RNA-seq

Total RNA quality was first assessed using the Agilent 2100 Bioanalyzer (Monash Genomics & Bioinformatics Platform). All samples showed high RNA integrity, with RIN values > 9.0. High-quality RNA samples were then submitted to BGI for library preparation and sequencing. RNA quality was re-checked at BGI, where all samples again met quality control criteria, with RIN values > 9.0 and 28S/18S rRNA ratios > 1.5. RNA sequencing was performed by BGI using the DNBSEQ platform (DNBseq™ Eukaryotic Strand-Specific Transcriptome Resequencing), generating 150 bp paired-end reads with approximately 20 million reads per sample. The experimental design included three biological conditions: wild-type (WT), knockout (KO), and heterozygote condition (KO/V84M), each with three independent biological replicates. Raw sequencing data were quality-checked using FastQC to assess read quality, GC content, and potential adapter contamination. Reads were aligned to the human reference genome (GRCh38.p14; NCBI GCF_000001405.40) using a pre-built genome index. Gene-level read counts were generated for downstream analysis. Differential gene expression analysis between groups was performed using the DESeq2 R package to identify significantly differentially expressed genes across comparisons.

### TOPFLASH/ FOPFLASH luciferase assay

TOPFLASH luciferase reporter contains TCF/LEF binding sites for beta-catenin. High Wnt/beta-catenin signalling activity results in beta-catenin binding to these sites, inducing luciferase expression measurable by using DUAL luciferase kit (Promega, USA, cat# E1910). FOPFLASH luciferase reporter has mutated nonfunctional TCF/LEF binding sites. Renilla luciferase-expressing plasmid (pRL-SV40, Promega, USA) was used to normalize transfection efficiencies.

HEK293T KO cells were seeded at the density of 50 000 cells/well onto 12-well plates. *MAP3K7* WT or MT constructs were co-transfected with either TOPFLASH and Renilla or FOPFLASH and Renilla using Lipofectamine 2000 (Invitrogen, cat# L3000015). p3xFLAG-CMV empty vector was used as a negative control. Cells harvested at 48 hours post-transfection were lysed and luciferase activity was measured on BMG LabTech ClarioStar system (BMG Labtech, Germany) following the manufacturer’s protocol. Beta catenin expression construct was used as a positive control. Firefly luciferase activity was normalized to *Renilla* luciferase activity for each transfected well. Values are the mean ± SEM of experimental triplicates from three independent transfections.

### Western blot

Cells were lysed using RIPA buffer supplemented with protease inhibitor cocktail tablets (complete™, EDTA-free, Roche, Cat N 04693132001) to preserve protein integrity. Lysates were incubated on ice for 30 minutes with occasional mixing and then centrifuged at 12,000– 14,000 × g for 30 minutes at 4 °C to remove cellular debris. The supernatant was collected, and protein concentration was determined using a bicinchoninic acid (BCA) assay (Pierce™ BCA Protein Assay Kits, ThermoFisher, USA, cat# 23225).

Equal amounts of protein (20–40 µg) were mixed with loading buffer (kit name), denatured at 95 °C for 5 minutes, and separated by SDS-PAGE using NuPAGE™ Bis-Tris polyacrylamide gels (4–12%, ThermoFisher, USA). Proteins were then transferred onto a PVDF membrane using a wet transfer system performed on ice. Following transfer, membranes were blocked in 5% skim milk prepared in TBST for 1 hour at room temperature to minimise non-specific binding. Membranes were subsequently incubated overnight at 4 °C with primary antibodies including p38 (Cell Signalling Technologies, cat# CST4511S) and p-p38 (Cell Signalling Technologies, cat# CST8690S), pTAK1 (Cell Signalling Technologies, cat# CST4536), and GAPDH (ThermoFisher Scientific, car# MA5-15738) diluted according to the manufacturer’s recommendations in blocking buffer. After washing with TBST, membranes were incubated with IRDye-conjugated secondary antibodies (IRDye 680 donkey anti-mouse IgG, LicorBio, cat# 926-68072 or IRDye 800 donkey anti-rabbit IgG, LicorBio, cat# 926-32213) diluted according to the manufacturer’s instructions, for 1 hour at 4 °C. Protein bands were visualised using a BioRad ChemiDoc imaging system under fluorescence detection settings. Protein expression levels were normalised to a GAPDH housekeeping protein. Semi quantification of protein expression band was performed using Image Lab software.

### Statistical analysis

Data was analyzed using GraphPrism9 software. Two-way ANOVA was used for multiple groups comparison and for calculation of p values. A p value < 0.05 was considered as statistically significant.

## Results

### Clinical report

#### Patient 1

A male phenotypic child presented with scoliosis, advanced bone age, high stature, camptodactyly, cleft palate, joint contracture, mild intellectual disability, speech delay, keloid scar and facial dysmorphisms including hypertelorism, large nasal bridge, epicanthal folds, retrognathia, and large ears. DSD features were micropenis, subcoronal hypospadias, small testis, bilateral cryptorchidism which was corrected. Hormone evaluation at first assessment showed low testosterone, high LH and FSH. MRI revealed periventricular hyperintensities and enlargement of the subarachnoid space. Initial clinical suspicous condition was overgrowth syndrome. The patient has 46,XY karyotype with *SRY* gene (+) by Fluorescent In Situ Hybridization (FISH). WES analysis identified a novel missense variant NM_145331.3 (MAP3K7):c.250G>A; p.V84M. None of rare variants on genes associated with overgrowth syndromes was identified. Taken together, FMD2 is likely a diagnosis for this patient.

#### Patient 2

A male phenotypic child born at 39 weeks. Pregnancy was complicated with mild growth restriction borderline hypoplastic aortic valve, and mitral valve. Prenatal diagnosis using amniotic cells with microarray confirmed 46,XY karyotype, and no detected copy number variant (CNV).

A postnatal echocardiogram confirmed a mildly hypoplastic aortic arch and isthmus, a functionally bicuspid. He has short stature (<0.01 percentile), his weight is 1.13 percentile, and his head circumference is 31.19%, fail to thrive, mild clinodactyly at the 5th finger, and one cafe-au-lait macule. Perineal hypospadias was noted at birth. Both testes were descended. Last audiology evaluation revealed the right hear had severe hearing loss and the left ear had moderate hearing loss. The skeletal examination showed narrowed disc spaces from T9 to T10, partial fusion of C5-C5. L1 showed a posterior fusion defect. There is carpal fusion involving the capitate and the hamate bilaterally. Brain MRI was normal. Ultrasound of kidney was normal. He had delayed expressive language but was on track with his motor milestones. Hormonal profile including GH, glucose, thyroid hormone was in normal range. He has one healthy brother. No other family history of note. A de novo c.574A>G variant of *MAP3K7* was identified by a trio WES analysis. Phenotype was consistent with CSCF.

#### Patient 3

A male phenotypic child presented with short stature, mild intellectual disability, facial dysmorphism including prominent forehead, hypertelorism, flat nasal bridge, retrognathia, keloid scar and large ears. DSD features were micropenis, hypospadias, corrected bilateral cryptorchidism, bifid scrotum, and hypoplastic scrotal skin.

The patient has 46,XY karyotype with SRY gene (+) by PCR. WES analysis identified a novel heterozygous missense variant of *MAP3K7* c.195A>G; p.I65M. Taken together, FMD2 is likely a diagnosis for this patient.

Other cases from literature searching: A total of 44 patients diagnosed with CSCF and FMD2 from previous published papers were reviewed, particularly examining if their DSD phenotypes reported. Among 8 affected males out of a total 23 patients with CSCF, 5 individuals also had DSD symptoms including either cryptorchidism, ambiguous genitalia, or small testis. Two out of ten affected males with FMD2 also presented with cryptorchidism. Of note, none of affected females with CSCF or FMD2 (n = 26) had DSD phenotypes (Figure 1C). Of note, we found one family with three affected members diagnosed with CSCF. Genetic test revealed all three had a heterozygous missense variant V50del, paternal inherited. However, only affected males were reported having small testis, no DSD feature was found in affected female [10]. Most syndromic 46,XY DSD had *MAP3K7* variants were located within the tyrosine/serine/threonine kinase domain (amino acid 1-157) (Figure 2B). In total, we have identified 10 of 21 affected male probands including our three new cases with DSD phenotypes.

**Figure 1:**
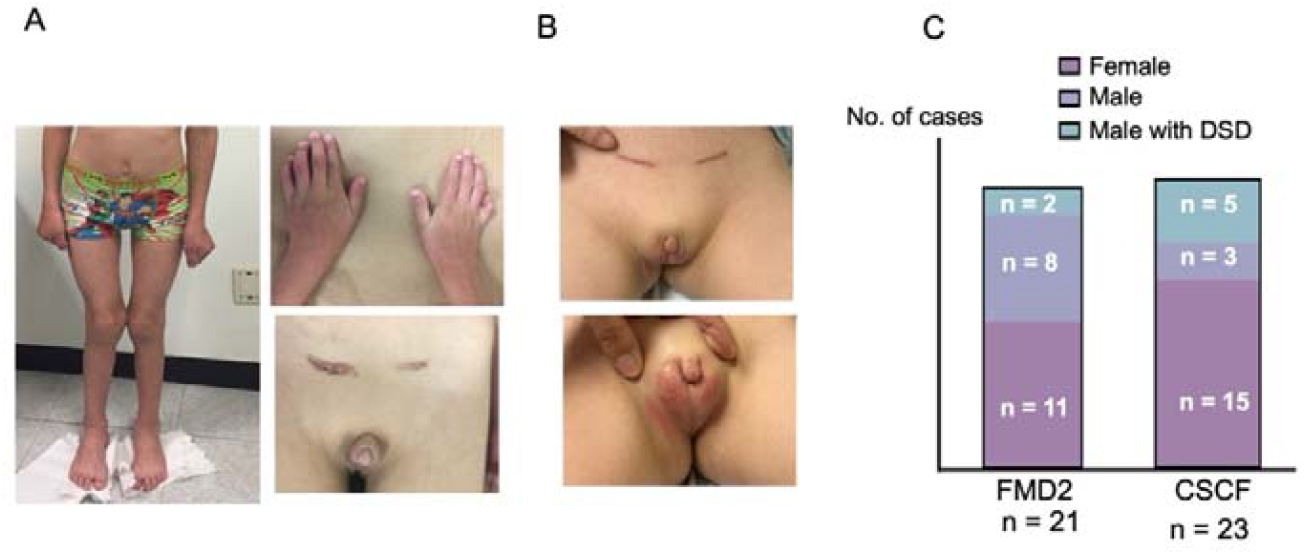
Clinical manifestation of patients carrying novel missense MAP3K7 V84M and I65M. A) Clinical phenotypes of patient 1 with V84M. B) Clinical phenotypes of patient 3 with I65M. C) Number of MAP3K7 cases with DSD from literature review. Patients consented for using their photos for publication.

**Figure 2:**
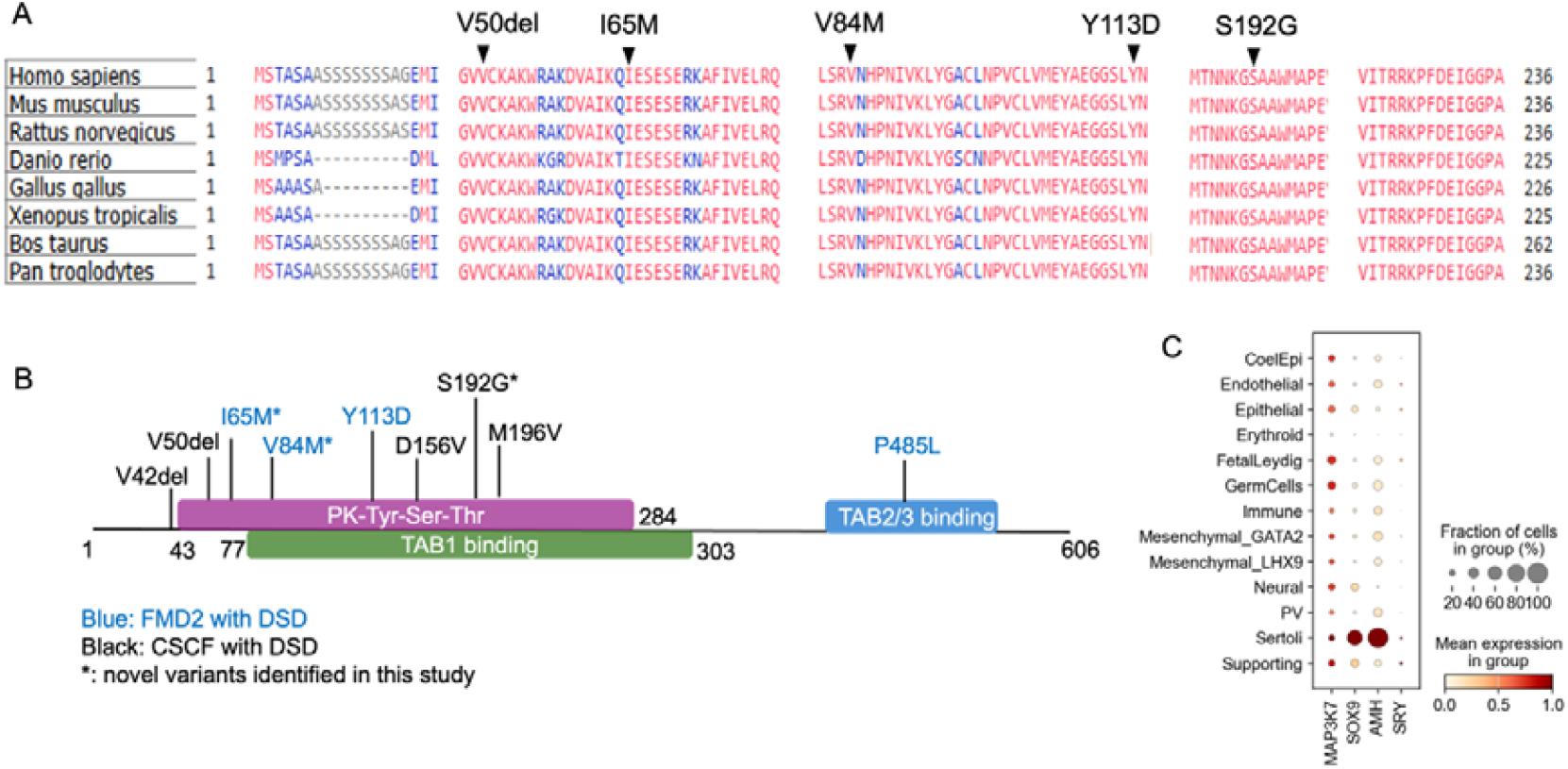
Characteristics of novel missense MAP3K7. A) Conservation of five investigated MAP3K7 variants. B) Schematic diagram of MAP3K7 structure. C) MAP3K7 expression in embryonic male gonads by scRNA-seq, achieved from [23].

### Prediction of deleterious variant effect and instability protein structure by in silico tools

All three missense variants identified in our new cases were absent in gnomAD v4 (https://gnomad.broadinstitute.org) and in ClinVar (https://www.ncbi.nlm.nih.gov/clinvar). AlphaMissense and CADD scores predicted deleterious effects for all three variants (Table 1). The positions of three novel missense variants were highly conserved across various vertebrate species (Figure 2A). I-Mutant predicted the three mutants decreased protein stabilities.

Next, we examined *MAP3K7* expression in human testis and ovary. Bulk RNAseq achieved from EMBL-EBI showed that *MAP3K7* is expressed higher in male embryonic testis than in female ovary (https://www.ebi.ac.uk). At single cell level, MAP3K7 is highly expressed in a subpopulation of Sertoli cells, Leydig cells, and germ cells (Figure 2C) (https://www.reproductivecellatlas.org/gonads/human-main-male/) [23].

### GATA4 and ZFPM2 downregulation in HEK293T cells lacking MAP3K7

*MAP3K7* is thought to cause CSCF syndrome via a loss of function mechanism while *MAP3K7* gain of function reportedly causes FMD2 syndrome. The association between MAP3K7 and 46,XY DSD has not been established, nor has the mechanism of wild type MAP3K7 function in male sex development. We generated *MAP3K7* deleted (referred to as MAP3K7^-/-^ or KO) and MAP3K7 V84M knock in in HEK293T cells using CRISPR-Cas 9 technique. For MAP3K7 KO in HEK293T cells, we inserted one nucleotide into exon 5, resulting to a frameshift mutation with the introduction of a premature termination codon. The sequence was confirmed by Sanger sequencing on DNA sample extracted from single clonal cell expansion. Consequently, no MAP3K7 protein was observed by Western blot analysis (Supp Figure1). We also introduced the V84M variant into one allele of MAP3K7 in HEK293T cells, the other allele had one nucleotide insertion, predicted to frameshift mutation which lead to no protein (referred to as MAP3K7^-/V84M^ or KI cells, Supp Figure1). The results were confirmed by Sanger sequencing and Western blot analysis as described above.

Three MAP3K7 KO clones and three MAP3K7 KI clones were expanded for RNA-seq and downstream functional experiments. Different genotypes were clustered by PCA plot (Figure 3A). A total of 2603 significantly differential expression genes (DEGs) were identified in KO cells, among which 1048 genes were downregulated and 1555 genes were upregulated. The number of DEGs genes in KI cells was 1588 including 721 downregulated genes and 867 upregulated genes (FDR<0.05). Notably, there were large numbers of overlapped DEG genes in the two datasets, either upregulated in both KO and KI cells or downregulated in both cell types. This suggests that disruption of MAP3K7 likely act via a loss of function mechanism (Figure 3B).

**Figure 3:**
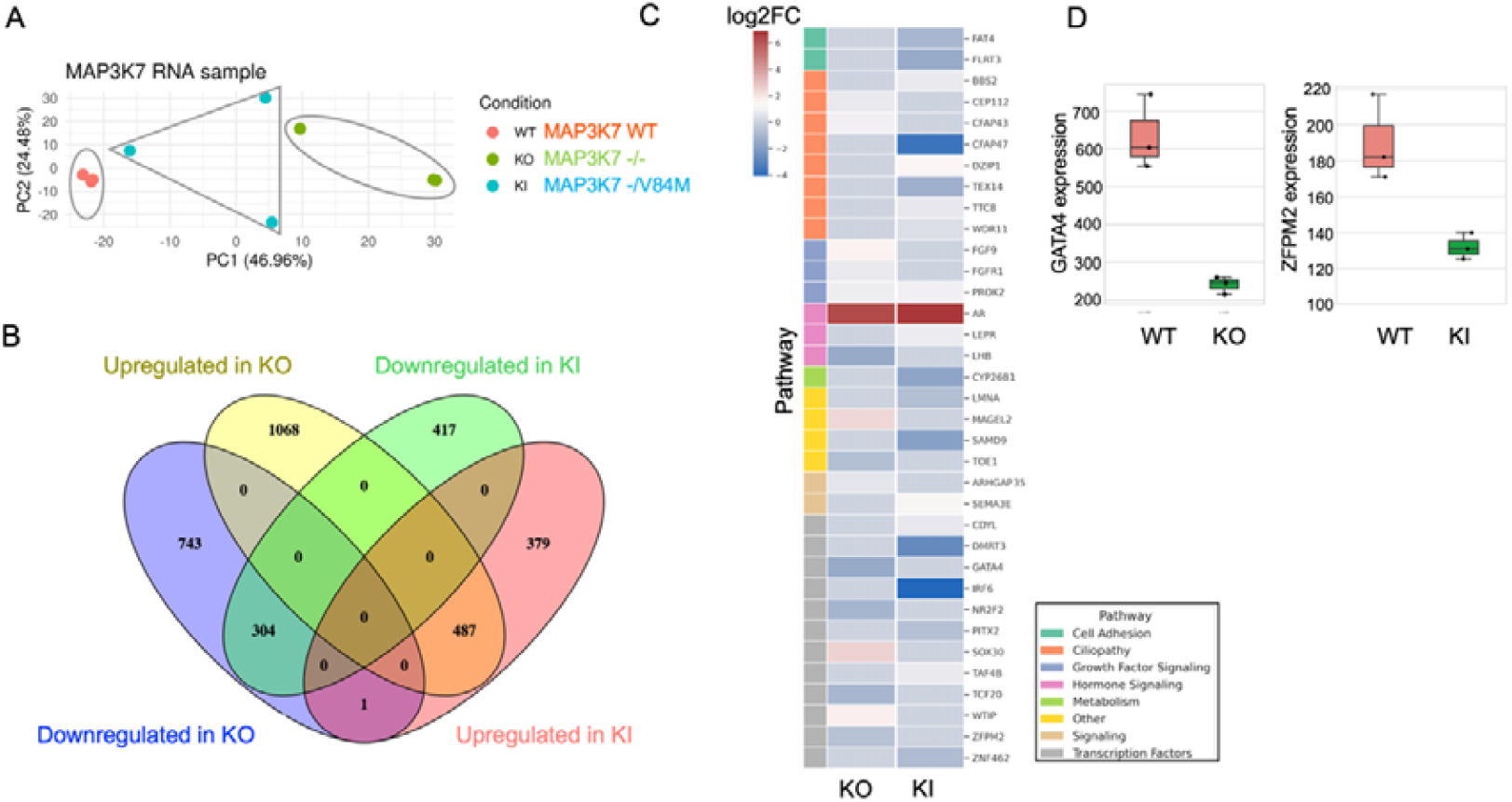
Differential expression gene analysis on HEK293T^MAP3K7-/-^ and HEK293T^MAP3K7-/V84M^. A) PCA results of samples subjected to RNA-seq analysis. B) Venn diagram of differential expression genes between HEK293T^MAP3K7-/-^ and HEK293T^MAP3K7-/V84M^. C) Heatmap of DSD genes identified in differential expression genes in the two datasets. D) Boxplot showing Downregulation of GATA4 and ZFPM2 in HEK293T^MAP3K7-/-^ cells. Expression of GATA4 and ZFPM2 was displayed as mean value of normalized read counts ± SD).

Next, we investigated DEGs for known DSD genes among in both KO and KI cells. There was 25 DSD genes found in DEGs of KO cells, and 20 DSD genes found in DEGs of KI cells, respectively. Of which, 11 DSD genes overlapped between the two datasets. These were *AR, BBS2, DZIP1, PROK2, SEMA3E, CDYL, FLRT3, WNT5A, PITX2, SAMD9, LMNA*. More than half of DSD genes involve in transcriptional regulation. Other related biological pathways include androgen hormonal signalling, ciliopathy, growth factor signalling, and cell adhesion (Figure 3C). We also found that GATA4 and its cofactor, ZFPM2, were downregulated in KO cells (Figure 3D). *GATA4* and *ZFPM2* plays an important role in male sex development as the GATA4-ZFPM2 complex upregulates *SRY* expression during sex determination. These findings suggest *MAP3K7* might be involved in male sex development by regulating the pro-testis pathway.

### MAP3K7 V84M, Y113D, S192G variants activate the beta catenin ovarian signalling pathway

MAP3K7 WT or mutant- (V50del, V84M, Y113D, I65M, S192G) overexpressing plasmids were co-transfected with TOPFLASH/ FOPFLASH luciferase reporter into HEK293T^MAP3K7-/-^ cells. After 48 hours of culturing, transfected cells were lysed and luciferase activity measured to determine whether the variants would affect the Wnt/beta catenin activity. Three MAP3K7 variants (V84M, Y113D, I65M) significantly activated beta catenin signalling pathway, of which V84M showed four-fold higher activation than MAP3K7 WT. In contrast, MAP3K7 V50del and S192G mutants reduced Wnt/beta catenin signalling when compared to MAP3K7 WT (Figure 4A).

**Figure 4:**
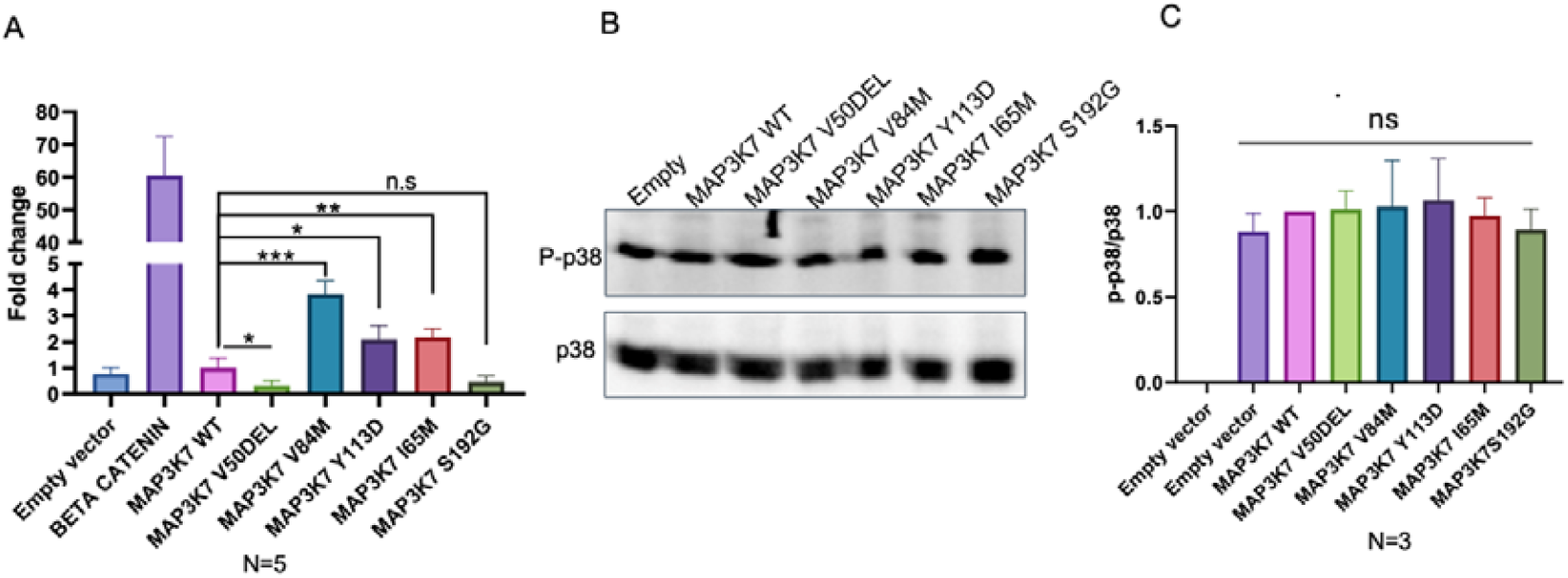
Results of experimental assays to validate the variant effects on Wnt/beta signalling pathway and phosphorylation of p38. A) TOPFLASH/ FOPFLASH result demonstrated V84M, Y113D, and I65M activated beta catenin signalling. B) Phosphorylated p38 (p-p38) expression by WB. C) Semi quantification of p-p38 and p38 showed all five variants had no effect on p-p38 level. The experiment was performed with three biological replicates. The values of mean ±SEM were displayed. * means p-value < 0.05; ** means p-value < 0.01; *** means p-value < 0.001. n.s means not significant

### MAP3K7 variants do not affect phosphorylation of p38

MAP3K7 WT or mutants (V50del, V84M, Y113D, I65M, S192G) were overexpressed in HEK293T^MAP3K7-/-^ cells. Protein was extracted from transfected cells and subjected to Western blot with Phospho-p38 and p38 antibodies to examine the phosphorylation level of p38, a downstream target of MAP3 kinase protein, and total expression of p38. WB results showed p38 and phosphorylated p38 were highly expressed in p3xFLAG-CMV empty vector, MAP3K7 WT and all five variants.

Semi quantification of P-p38 and p38 showed that none of *MAP3K7* variants affected the phosphorylation level of p38 (Figure 4B, 4C).

## Discussion

### Clinical manifestation and DSD

Phenotypically, the clinical manifestations in our patients are consistent with what had been described for CSCF or FMD2, autosomal conditions caused by MAP3K7 variants. Patient 1 and patient 3 are likely diagnosed with FMD2 given the predominant abnormalities of skeletal system, keloid scar and absence of cardiac defects. Patient 3 with the most severe phenotypes likely has CSCF as he failed to thrive, had hypoplastic aortic arch, isthmus and hearing loss.

We identified three novel MAP3K7 missense variants (V84M, I65M, S192G) in three patients by WES. Only variant S192G is confirmed *de novo* by trio WES. The other two were uninformative due to unavailability of parental DNA samples. All three variants are novel since absent in gnomAD v4 and ClinVar databases. The in silico tools predicted deleterious effects for three variants by CADD score and AlphaMissense, warranting functional studies. Pathogenic missense variants are the most common type in CSCF and FMD2.

Initially, patient 1was suspected to have overgrowth syndrome as his height exceeded the 99^th^ percentile for age-matched male children. By WES, no variants were found in know overgrowth associated genes. Instead, we identified a missense variant V84M in *MAP3K7* which could explain the CSCF/FMD2 syndromic manifestations in this patient, but not his tall stature. In CSCF or FMD2, normal height or short stature is commonly seen.

Among the three new affected individuals, patient 3 with the I65M *MAP3K7* variant shows the least severe clinical features, with minor skeletal anomalies. Subjects with substitutions in the MAP3K7 kinase domain have a notably milder phenotypes than those with recurrent P485L variant [11, 24]. Variants causing FMD2 increase MAP3K7 autophosphorylation, suggesting a gain of function mechanism; conversely variants causing CSCF decrease MAP3K7 autophosphorylation resulting in dysregulation of downstream signalling pathways [10, 11, 25]. This previous data suggest an optimum window of kinase activity outside which clinical disparities between the two syndromes occur.

The presence of DSD symptoms including cryptorchidism, hypospadias, micropenis, and small testis in our male patients is a striking finding. Through intensive literature searching, we also found other male cases described with either cryptorchidism, ambiguous genitalia or small testis. Of note, majority of cases with DSD had *MAP3K7* variants locating in the N-terminal kinase domain. No affected females had DSD features (Figure 1C, 2B). This strongly suggests that *MAP3K7* plays a role in human male sex development.

MAP3K1, another MAP3 kinase member, is a known causative gene of 46,XY gonadal dysgenesis and sex reversal. P38 is linked to male sex development as a downstream target of MAP3K1 in human or Map3k4 in mice. Ostrer et al. proposed a model where MAP3K1 disruption acted via gain of function, increased phosphorylated p38, resulting to tilting the balance from SOX9/FGF9 driving testicular development to WNT/beta catenin signalling critical for female sex development [26, 27]. In contrast, Greenfield et al. showed that Map3k4 loss of function inhibiting mouse testis determination via p38 MAPK-mediated control of Sry expression[28]. By accessing scRNA-seq of human male gonad tissues [23], we found that MAP3K7 was highly expressed in a subset cell populations in Sertoli cells, Leydig cells, and other cell types (Figure 2C).

Our studies suggest that MAP3K7 behaviour differs from MAP3K1. MAP3K7 KO cells downregulated expression of *GATA4* and *ZFPM2*, two essential regulators of SRY during testis development. Overexpression *MAP3K7* variants V84M, Y113D, I65M activated beta catenin signalling, a pro-ovarian function. Therefore, *MAP3K7* mutations might cause DSD by weakening testicular pathway and/or activating Wnt/beta catenin pathway, but not via p-p38.

## Conclusion

We present here three new cases with syndromic 46,XY DSD carrying three novel missense variants located in the kinase domain of *MAP3K7*. This study is the first to draw a link between MAP3K7 and DSD, showing functional effects in 3 cases and identifying 7 earlier cases of FMD2/CSCF with DSD symptoms. Functional studies in vitro suggest MAP3K7 might involve in male sex development by promoting testicular pathway and suppressing Wnt/ beta signalling pathway. MAP3K7 disruption can cause DSD via loss of function mechanism.

**Supplemental Figure 1:**
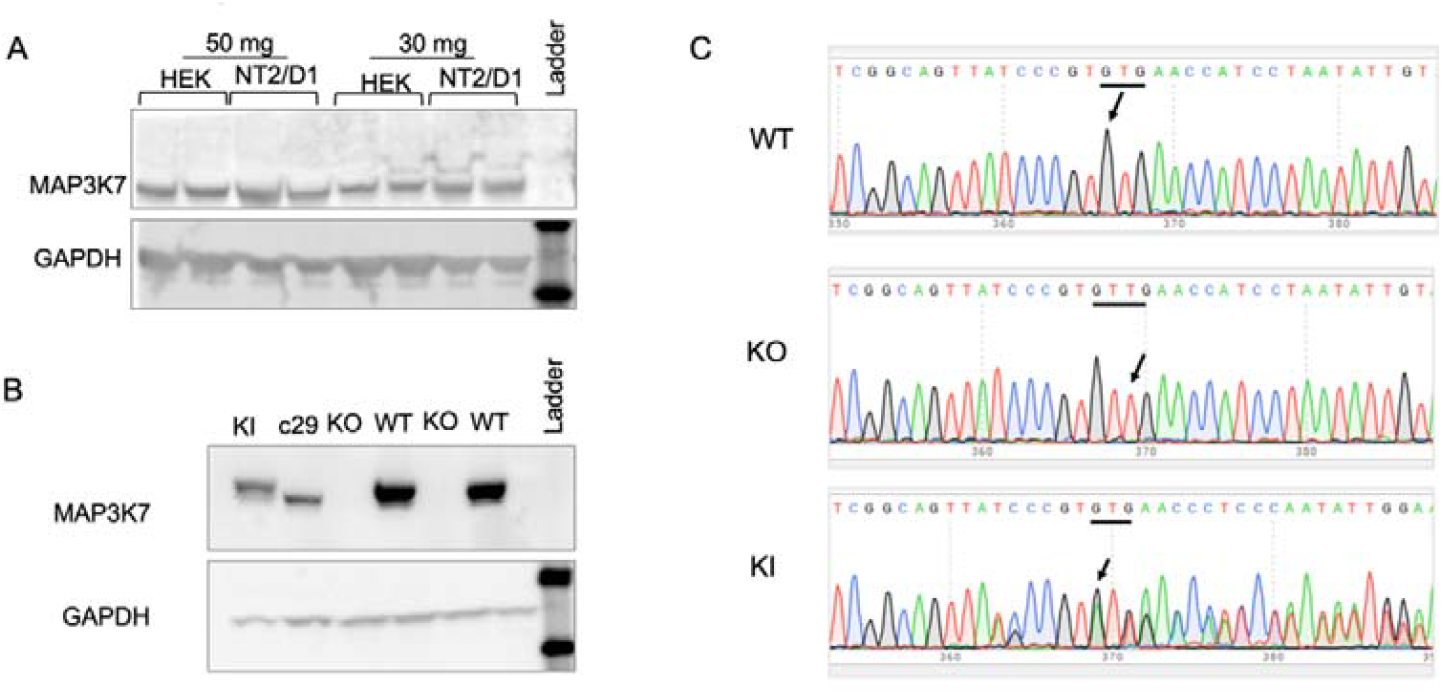
WB and Sanger sequencing results of endogenous MAP3K7 and CRISPR KO/ V84M KI in cell lines. A) MAP3K7 endogenous expression in HEK293T and NT2/D1 cells by WB. B) Complete MAP3K7 KO and half dose expression of V84M KI confirmed by WB. C) Sanger sequencing confirmed 1 bp insertion leading to a frameshift mutation, nonsense mediated decay predicted, and null protein as shown in B); and heterozygous V84M KI with the other KO allele.

**Supplemental Table 1:**
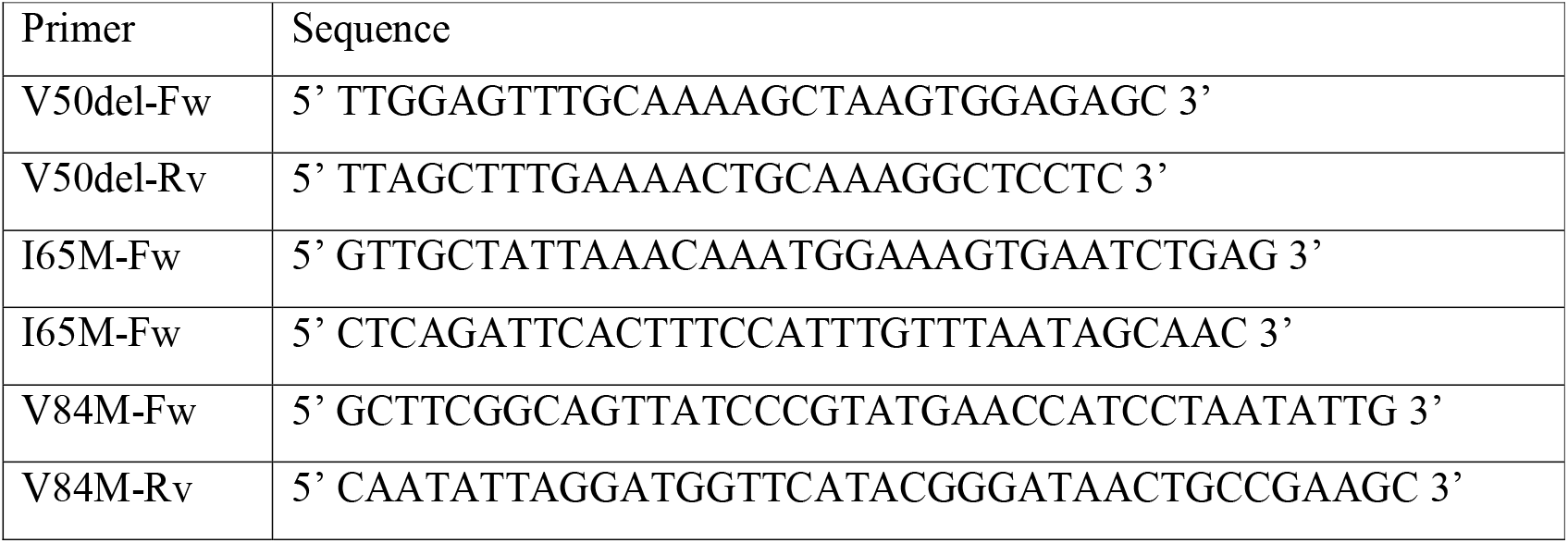
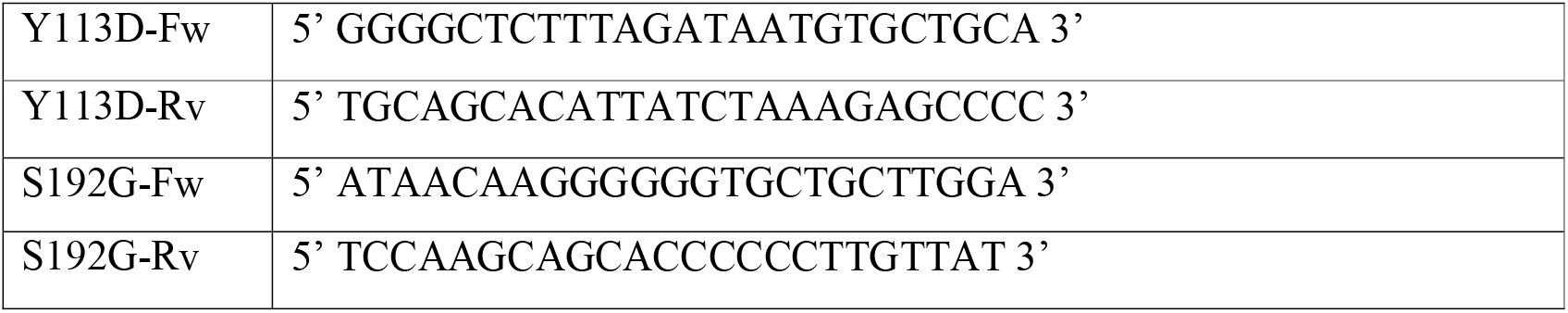
Primers used for mutagenesis.

## Data Availability

All data produced in the present study are available upon reasonable request to the authors

## Acknowledgements

The authors would like to thank the patients and their families participating in this study. We thank Prof. Stephen Robertson for the kind gift of MAP3K7 constructs. We thank Prof. Ken McElreavey at Pasteur Institute, France and Prof. Eric Vilain, UCLA, USA and Dr. Emmanuele Delot, Washington University, USA for sharing MAP3K7 variant findings in their DSD patient cohorts.

